# Exploring rat meat consumption patterns, and perception of risks regarding urban rats; implications for rat-borne zoonoses outbreaks and drug resistant pathogens spread in urban areas of Ghana

**DOI:** 10.1101/2024.04.23.24306236

**Authors:** Godwin Deku, Emmanuel Younge, Stephen L. Doggett, Rofela Combey, Isaac Kwame Badu, Mercy Amo Paintsil, Smile Kwabena Ametsi, Angela Ama Wills, Rabiatu Bonnoua Bonney, Kwabena Owusu Amoah

## Abstract

**Introduction:** This research explores rat meat consumption patterns among urban Ghanaians and their perception of risks associated with urban rats. Two hypotheses on risk perception among urban inhabitants were proposed: the risk of contracting diseases by consuming rat meat and the risk of contracting diseases from peridomestic rats.

**Method/Findings:** To achieve the objective, a descriptive cross-sectional survey using a questionnaire was conducted that recruited participants from urban settings in Ghana. Of the 829 respondents recruited, around 65% consumed rodents, and of these, 35% consumes rat meat. Through statistical analyses, our data revealed gender, age, region, religion, ethnicity, education, profession and income significantly influenced rat meat consumption and residents’ perception of disease risk. An adjusted multivariable model identified males aged 27 to 34years with no formal or a basic education in southern Ghana as the most likely rat meat consumers. The majority of the participants (60.3%) indicated rats are often present in homes and community drains, and have some awareness where rats can be found in their place of residence. Low perception scores regarding disease risks associated with rat meat consumption were recorded, with the majority of respondents (55-74.9% of 829) perceiving that there were minimal disease threats associated with peridomestic rats. The outcome of this belief was that participants undertook limited rat control in their neighborhood.

**Conclusion:** The poor perceptions of disease risks regarding rats increases the vulnerabilities of urban Ghanaians to zoonotic spillovers. This underscores the critical need for public education on rat-borne zoonoses in urban Ghana. This survey presents the first baseline study on urban inhabitants’ interactions with peridomestic rats in Ghana and the data will be crucial in the on-going interventions by the Ghana Health Service to minimize transmission of Lassa fever and other rodent-borne diseases and the spread of rodent related drug resistant pathogens.

**Author Summary:** Human engagement with rats in urban settings provides a mechanism of transferring rat-related zoonoses to the human population. Unlike some other African nations, instances of zoonoses relating to rats are not common in Ghana. Consequently, the public’s perceptions of disease threats posed by urban rats remain largely unexplored. Ghana’s Lassa fever problems began in 2012, and in 2023, the country experienced 14 cases with one death. The study herein enhances our understanding of the epidemiological risk factors in evaluating Ghanaians preparedness against rat-borne zoonoses in urban settings, by examining rat meat consumption patterns and associated risk perception with the rats. A total of 829 Ghanaians were interviewed from urban residential areas on rodent risk behaviors. Results revealed that around 35% of people consume rat meat in urban settings of Ghana and low perception scores on disease risks pertaining to the rats were recorded. The low scores are reflected in the limited attempts by the public to control rats. Rat meat consumption and perception of risks were driven by several sociodemographic variables. Our data could be used by the Ghana Health Service to justify implementation measures for rodent management to mitigate Lassa fever and the spread of antimicrobial resistant pathogens.

## Introduction

Global population growth and rapid urbanization poses a critical economic and health challenges [1,2]. This has correlated with the escalation of urban pest populations, including disease vectors, leading to the emergence of new vector-borne illnesses [2,3]. The United Nations estimated that 4.2 billion people currently reside in urban areas worldwide with projections indicating that by 2030, 75% of the global population will live in urban areas [4]. Even Africa and Asia, the least urbanized continents, are anticipated to witness substantial urbanization by 2050 [5] with Ghana having over 50% of its population currently residing in urban areas [6]. As population growth drives expansion of urban land cover, estimated to increase by 1.2 million km^2^, biodiversity loss becomes inevitable to sustain human population density [7]. Consequently, a significant portion of the world’s population is expected to coexist with urban pests in the foreseeable future [8] and human-pest interactions may increase if control measures are not implemented [9]. Urbanization facilitates pest proliferation due to factors such as industrialization, agricultural and infrastructural development, habitat degradation, forest decline, excessive waste generation, and poor environmental management [10,11]. Rats (*Rattus* sp.), classified as small mammals within the order Rodentia, inhabit forests, peri-urban and urban areas [12]. Their biology, behavior, ecology and adaptability to human environments have been extensively studied due to their economic, medical, social and scientific importance [12,13,14,15,16]. Of the 130 species in the Rattus genus, *Rattus norvegicus* (Norway rat), *Rattus rattus* (black rats) and *Rattus. tanezumi* have successfully colonized urban ecosystems throughout much of human civilization [17,18]. The Norway rat also called the brown rat or the sewer rat is distinguishable from the black rat by its tail, colour, ears and by other morphophysiological features [17]. It exhibits superior competitive aggressiveness and has become one of the most dominant mammals globally [19].The rise in global rat population can be attributed to shifts in human environment, ecology and climate ([2,20] with research indicating a heightened risk of rat infestation in neighborhoods characterized by high poverty levels, aged buildings, dense housing systems and inadequate sanitation [11,21].

Rat meat serves as an important protein source in many countries particularly across Africa, Asia and the Middle East [22,23,24,25,26]. Research in Guinea revealed that 91.5% of the population highly value protein from rodent sources [24]. In Nigeria, of the 200 and 155 study respondents interviewed from urban and peri-urban neighborhoods, 33.5% and 31.1% respectively, consume urban rodents [23]. Various rodent species are consumed in Ghana [27,28]. However, over the past six decades, the majority of the 300+ newly identified infectious diseases have stemmed from wildlife with human-wildlife contact serving as a major transmission pathway [29]. Rodents alone provides reservoir of an estimated 50% of emerging zoonotic pathogens and commensal rats are an ecosystem of numerous disease agents with over fifty-three infectious agents identified thus far [3,20,30,31,32,33]. Diseases such as toxoplasmosis, salmonellosis, plague, leptospirosis, rat-bit fever, hemorrhagic fevers, and zoonotic babesiosis have been associated with rats [3,33]. Transmission of these disease pathogens to humans occurs through various means, including via contamination of water and food, and contact with rat feces and urine [3]. Consuming rat meat raises a more critical risk facilitating the transmission of zoonotic pathogens and epizootics from rat populations to densely populated humans. The act of inhaling aerosolized pathogens from rat fur, the potential exposure to blood-borne and saliva related infections during the carcass handling and butchering, as well as the consumption of undercooked meats, further heighten the risk of zoonotic transmission [34]. These risks were underscored by studies such as Douno and colleagues [35], which explore hunting and consumption of rodents and Mangessho and Colleagues [36] which examine knowledge and perception of pastoralists in Tanzania and hunting and consumption of peridomestic rodents and disease transmission risks in relation to this issue were also discussed by Ter Meulen and colleagues [24]

Regions such as West and East Africa have witnessed a prevalence of rat-borne diseases, with Madagascar experiencing frequent plague outbreaks in recent decades [37,38,39,40,41]. Lassa Fever, endemic in most African countries, poses a more critical threat, primarily transmitted by rats in the genus *Mastomys* [42]. Among the major zoonoses in West Africa, Lassa Fever causes approximately 100,000-300,000 infections with around 5,000 deaths annually [42,43], Transmission of the disease may occur through contact with rat feces and body fluids [43,44]. Several studies have demonstrated a correlation between increases in rat populations and the proliferation of pathogens vectored by rats [20,45,46]. This escalates the risk of epizootic events spilling over into human communities, highlighting the importance of human-rat interactions. Urban rats inhabits diverse habitat types that are under high chemical/drug selection pressures for pathogens and include waste disposal sites, livestock yard, urban agriculture sites, municipal waste water systems, and biomedical/health care/hospital wastes depots which all harbor a wide range of pathogens [47]. These pathogens may be spread to community food sources, water bodies, household insects, and domestic animals including poultry, and pets through commensal rat activities [47,48]. Drug resistant *Staphylococcus aureaus* were found in household soil, rodents and chickens in Tanzania [49]. In a recently published study, the common rats; *R. norvegicus* and its cousin *R. rattus* were identified in mediating the transfer of multiple drug resistant *Enterobacteriaciae* from livestock into human environment [48]. In this study, 28 of the 53 *E. coli* strains and two of the five *Salmonella* strains identified were multiple drug resistant suggesting urban may be playing a crucial role in the dissemination of antimicrobial resistant pathogens (AMR) [31,32,47,48,49,50,51].

Reducing rat populations, particularly in urban settings, presents significant challenges due to various factors. These include dense human populations, extensive infrastructural development [52], nocturnal and sub-nocturnal rodent behavior, the opportunistic nature of urban rats, and their adeptness at establishing nests in obscure locations within human dwellings, and their prolific reproductive ability. Moreover, the behavioral resistance of rats to toxic baits [53] and the resurgence of pesticide resistant rat populations, the banning of glue traps, and second generation anticoagulants, have further exacerbated the global challenge of rat control [54]. In 2020, the Ghanaian media reported a significant event where approximately ten thousand urban rats were killed overnight in the capital city following a major pesticide incident during the active phase of the COVID-19 pandemic, which was indicative that urban rats’ in the capital city have greatly increased. The escalating density, particularly in public places implies an augmented risk of rat-related pathogen transmission to the Ghanaian populace. Despite the cohabitation of Ghanaian urban residents with commensal rats, scant literature addresses the perceptions and apprehensions of the community regarding these rodents. To the best of our knowledge, no prior assessment has been conducted to characterize commensal rats in peridomestic and urban areas in Ghana and the associated potential disease risks. Additionally, public knowledge, perception and awareness of the health risks of rats remain largely unexplored. Previous research in a rural area in Ghana and most countries in Africa and Asia suggested poor knowledge and perceptional unawareness on the disease risks associated with wild animals [55,56,57,58]. This research herein aims to provide a comprehensive overview of the Ghanaian public’s rat meat consumption patterns and their perceptions regarding urban rats. We consider two pathways as determinants for potential zoonotic outbreaks: rat meat consumption and human interaction with rats in the urban environment. This paper examines the potential risk for rodent-borne diseases in urban settings of Ghana.

## Materials and Methods

### Ethical Clearance

The study received ethical review and approval from the research and publication committee of Accra School of Hygiene under the Ministry of Sanitation and Water Resources affiliated to the University of Cape Coast, Ghana (Approval number: SOH/MSWR/06/01/2023/001). Voluntary anonymous respondents were informed on their willingness to form part of the survey. None of our study participants is below the age of 18(yrs.) and all participants provided informed oral consent before the interview. Written consent were not obtained because not all our study participants were literate hence, the facilitator approved the consent note on their behalf after seeking for their consent response.

### Study area

According to the recent population Census [6], Ghana’s population has increased by approximately six million since 2010, reaching a current total population of over 30.8 million in 2021. Administratively, the country is divided into 16 regions (Fig 1) comprising 261 districts. Seven are urbanized and currently over 17 million of the total population is now residing in urban areas. Southern Ghana has eleven (11) regions whereas, the North has five (5). The country has two major religious groups with Christianity being the predominant religion followed by Islam and traditional beliefs. Islam is more prevalent in the northern region of Ghana. Ghana is linguistically diverse, some 79 ethnic groups speaking over 40 languages and 75 dialects [59] with English serving as the national language. The five largest ethnic groups are Akans, Mole-Dagbon, Ewes, Guans, and the Ga-Adangbe. The Akan ethnic group include subgroups such as Asante, Fantes, Akuapim, Akropong, and Akyems constitute around half (47.5%) of the nation’s population and is mainly distributed in the south. Northern Ghana is dominated by the Mole-Dagbon ethnic group and include the Dagombas majority, Mossi, Mamprusi, Nanumbas and Gonja ethnic group comprising 16.6% of the nation’s population [59]. The remaining ethnic groups, including the Ewe, Ga and Guans constitute about 35.9% of the population [59].

**Figure 1.**
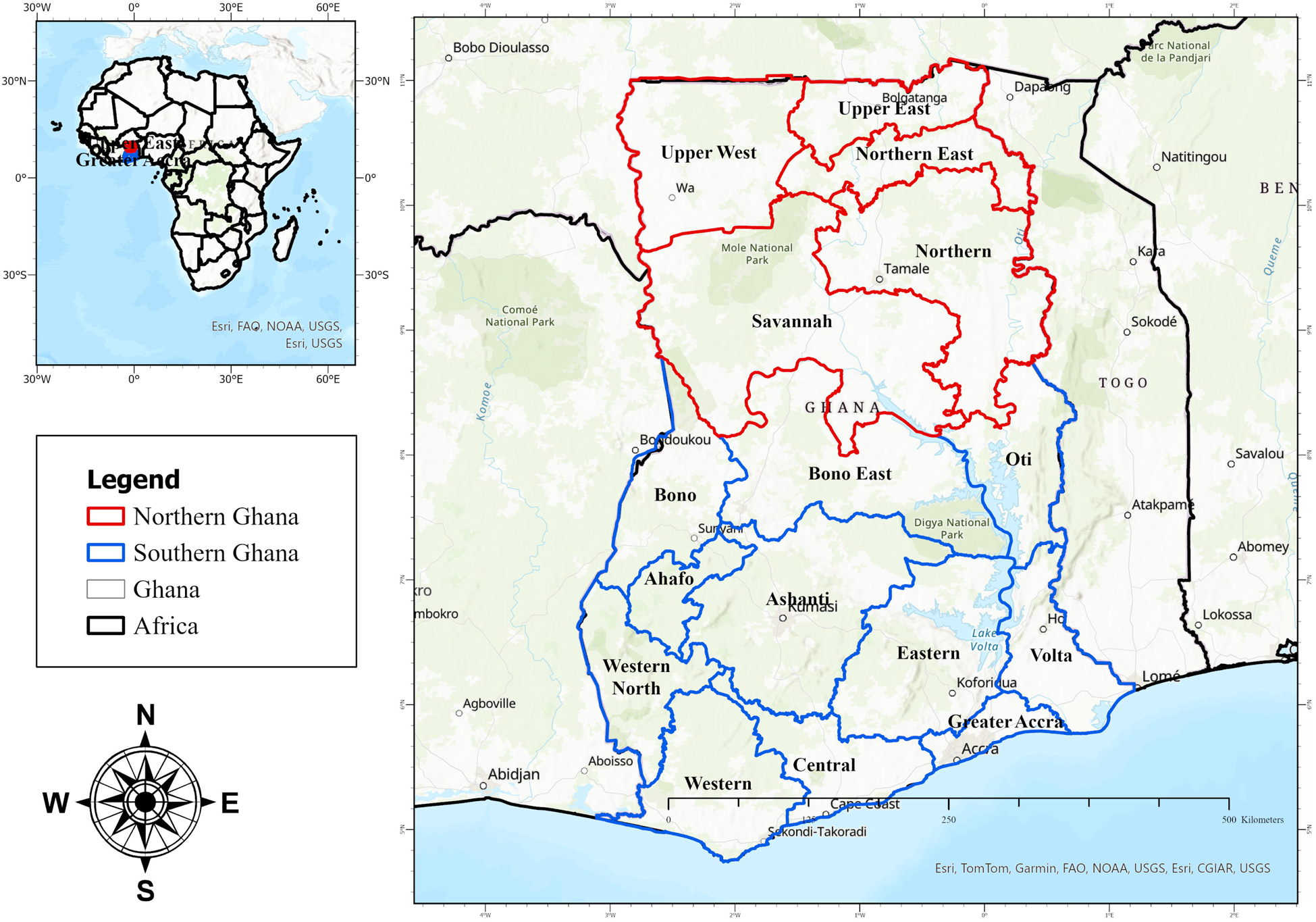
Map of Ghana showing the 16 administrative regions in the north and southern divisions.

### Study design and data collection instrument

This study utilized a descriptive cross-sectional survey aimed at providing a general overview of the rat meat consumption rate among the Ghanaian public, as well as their perception of associated risks, and the sociodemographic drivers. Data collection was conducted solely through interviewer-administered structured questionnaires, predominantly comprising closed ended questions. Respondents were interviewed and guided by a facilitator to complete the questionnaire, with the process typically taking around 10-15mins per participant. The questionnaire design drew upon previous publications [23,34,56,60,61]. Prior to implementation, the questionnaire was pre-tested on 20 respondents in the capital city, Accra Ghana and was subsequently converted to Google forms and shared among interviewers.

### Data collection

Environmental health officers and environmental health trainees collected the data for the study. The questionnaire links were sent to these groups via email, and WhatApps to individual Environmental health officers and trainees in the 261 districts located in the 16 administrative regions in Ghana. The questionnaire provided clear instructions and guidelines at the outset, accompanied by the inclusion of a brown rat logo to facilitate recognition by participants. Interviewing Officials who required clarification utilized a phone number provided in the questionnaire. In collecting the data, the officers and trainees firstly displayed the rat picture logo featuring the popular rat local name ‘ekusie’ to aid in recognition by the participants. Interviews were conducted exclusively with individuals residing in urban areas. Participants were anonymous randomly approached at various locations such as streets, lorry stations, shops, hospitals neighbourhood, schools, and other meeting points. Upon consenting, participants were given the questionnaire by the interviewer. Completed questionnaires were submitted to the authors online by the official interviewers. Personal information such as participants’ names, and phone numbers were not collected to maintain confidentiality. The survey was conducted over a period of five months from September 2023 to January 2024 and inclusion criteria was based on the respondents who resided in an urban area of Ghana. Participant from rural areas/villages and unknown areas were excluded. Only participants above the age of 18years were interviewed.

The questionnaire was divided into five sections. The first section collected data on socio-demographic characteristics of the study respondents. Data such as region of the respondent, gender, ethnicity, age, education, profession, monthly income, and marital status were recorded. The second section of the questionnaire which included mainly ‘Yes or No’ responses, collected data on rat meat consumption. This included information on whether participants were familiar with rodents and if they consume rodent/rat meats and whether they have different preferences for town rats or wild rats/bush rats, and if willingly purchased rat sold in restaurants. The third section addressed perceptions about health risks associated with rat meat consumption, including if participants perceive that rat carried disease agents and if they believe people could contract diseases from consuming rats. The fourth section collected data on people’s awareness on rat populations in their neighbourhood, such as if they have sighted rats in recent times or observed any rise in the rodent population within their neighborhood. The factors accounting for the rodent increases were recorded along with what they do if they detect rats in their premises. The final section of the questionnaire addressed opinions on the rat population, such as if respondents were concerned about rats, their reasons for the concern, and if attempts were made to manage rats in their neighborhood and homes.

### Data processing and statistical analysis

The questionnaire data was exported into Microsoft Excel version 2010 as an Excel file. The data was processed and checked for completeness and consistency. The cleaned data was imported into SPSS v26 (IBM Corp., Armonk, NY) for analysis. Descriptive statistics were presented in frequency and percentage proportions. The analyses of the study data utilized Independent Pearson’s Chi-square test and binary logistic regression (logit) models to demonstrate association between independent variables/predictors (sociodemographic variables) and dependent variables (Yes/No responses). Odd ratios and their corresponding 95% confidence intervals were determined from the logistic models. Perception is a latent construct and perception responses were measured using Likert scale. The Likert scale employed five optional responses which included, agree strongly, agree, neutral, disagree and disagree strongly. The highest correct perception score response was ranked/assigned 0.5, and 0.3 for neutral and 0.1 for every incorrect/lowest perception score about a statement. Mean perception scores were compared using non-parametric tests. Mann Whitney U test was used to test the significant proportional differences between two independent variables and the Likert responses (mean perception scores). Whereas, Kruskal Wallis tests of independence were used to compare significant proportional differences among three or more independent variables/predictors and their Likert responses. Pairwise post hoc analysis was employed with Bonferroni correction for multiple tests to separate means and the adjusted p-values were recorded. All assumptions behind the statistical parameters and fitness tests were met.

## Results

### Sociodemographic characteristics of the general public respondents

Table 1 depicts the sociodemographic profile of the respondents. In all, 829 questionnaires were received from 79 districts spanning 16 administrative regions throughout Ghana. The study encompassed 28 out of the 75 ethnic groups in Ghana with the Akans, Ewes, Gas and Mole-Dagbon constituting the predominant groups. The Akans and Mole-Dagbon, the primary ethnic entities in Ghana comprised the majority (61%) of the study participants. A higher proportion of responses originated from southern Ghana (65.7%) compared with the north. The gender distribution skewed towards males (59.8%). About half (53.2%) of the participants were unmarried. The age distribution revealed a significant majority (79.1%) falling within the 21-44 years group. A substantial proportion (63.6%) of the participants had attained senior high school education, or higher. Regarding religious affiliation, Christians accounted for more than half (53.4%). In terms of profession, students and traders constituted around 44.6%, while health professionals, teachers and others comprised about 38.3%. Nearly half of the study participants were not engaged in any form of employment. Regarding income 30.2% of the survey respondents reported earning between 300-500 Ghanaian cedis monthly whiles 27.9% reported incomes ranging from 600-1100 Ghanaian cedis monthly. Additionally, around half of the study participants resided in family-owned homes.

**Table 1.** Sociodemographic information of the study respondents.

| Sociodemographic variables | N (%) |
| --- | --- |
| **Gender** |  |
| Male | 496 (59.8) |
| Female | 333 (40.2) |
| **Marital Status** |  |
| Single | 441 (53.2) |
| Married | 333 (40.2) |
| Married in the past | 55 (6.6) |
| **Age** |  |
| <21 | 63 (7.6) |
| 21-26 | 245 (29.6) |
| 27-34 | 232 (28.0) |
| 35-44 | 178 (21.5) |
| 45-54 | 79 (9.5) |
| 55+ | 32 (3.9) |
| **Education level** |  |
| No formal edu. /Primary | 200 (24.1) |
| Junior High School | 102 (12.3) |
| Senior High School | 325 (39.2) |
| Bachelor and above | 202 (24.4) |
| **Regional Zones** |  |
| Northern Ghana | 284 (34.3) |
| Southern Ghana | 545 (65.7) |
| **Religion** |  |
| Christianity | 443 (53.4) |
| Islamic | 324 (39.4) |
| Traditionalist/Atheist/Others | 59 (7.0) |
| No data | 3 (0.4) |
| **Ethnicity** |  |
| Dagomba | 253 (30.6) |
| Akan | 252 (30.4) |
| Ga | 104 (12.5) |
| Ewes | 81 (9.8) |
| Mamprusi | 22 (2.7) |
| Dagati | 20 (2.4) |
| Frafra | 11 (1.3) |
| Guan | 5 (0.6) |
| Others | 82 (9.9) |
| **Profession** |  |
| Students | 202 (24.5) |
| Health Professionals/Teachers | 165 (19.9) |
| Traders | 224 (27.1) |
| Others | 234 (28.5) |
| No data | 4 (0.5) |
| **Employment status** |  |
| Not employed | 385 (46.7) |
| Employed | 436 (52.8) |
| Retire | 4 (0.5) |
| No data | 4 (0.5) |
| **Income (GHS.) (\$1= GHS. 13)** | |
| <500/No income | 254 (30.6) |
| 600-1100 | 231 (27.9) |
| 1200+ | 145 (17.7) |
| No data | 199 (24.0) |
| **Residential status** |  |
| Own the home | 155 (18.8) |
| Family home | 440 (53.3) |
| Renting | 216 (26.1) |
| Not classified | 15 (1.8) |

### Rat meat consumption

Table 2 presents the findings on the participants’ consumption practices. Out of the 829 participants, the high majority (82.2%) exhibited familiarity with rodents and most (64.8%) acknowledged consuming such animals. However approximately 35% of those who consumed rodents consume rat meat. Around 22% had consumed rat meat recently and about 34% expressed willingness to consume the meat when presented with the opportunity. On average, 26% of the respondents could recall someone who had consumed rat meat recently, expressed willingness to buy rat killed from their neighbourhood, consumed rat meat sold in a restaurant or engage in hunting and consuming rats that visited their premises.

**Table 2.** Rat meat consumption among the respondents.

| Statement/Question | Response |  |
| --- | --- | --- |
|  | Yes (%) | No (%) |
| Familiar with rodents (such as grass cutter, rat and mice) | 698 (82.2) | 125 (15.1) |
| Consumes rodent meat. | 537 (64.8) | 288 (34.7) |
| Consumes rat meat. | 290 (35.0) | 535 (64.5) |
| Ate rat meat recently. | 184 (22.2) | 636 (76.7) |
| Willing to consume rat meat when presented with one today. | 278 (33.9) | 542 (66.1) |
| Remembered someone that consumed rat meat recently. | 204 (24.7) | 619 (74.9) |
| Willing to buy rat killed from the neighbourhood and sold at cheap price. | 202 (24.4) | 616 (74.3) |
| Willing to buy town rat meat sold in a restaurant/chop bar. | 198 (23.9) | 626 (75.5) |
| Willing to buy bush rat meat sold in a restaurant/chop bar | 234 (28.2) | 400 (48.3) |
| Willing to kill and consume rat that has visited your home. | 206 (25.0) | 623 (75.0) |
| **Average** | **26.04%** |  |

### Sociodemographic predictors of rat meat consumption

Table 3 presents frequency and percentage summaries demonstrating that a higher proportion of males (23.8% of 829) consume rat meat than females (11.4%). Additionally unmarried (singles) consume rat meat more than those who are married. Those in age 27-34 years consume rat meat more than the other age groups. Ethnically, Ewes (n=47 > 34), and the Mamprusi (n=15>7), significantly surpass non-rat meat consumers. Those with senior high school education, residing in the southern region part of the country, Christians and Akans, unemployed, and people having an income of less than 500 Ghanaian cedis, monthly, demonstrated a higher proportion of rat meat consumption than their counterparts. Thus there were associations between rat meat consumption and sociodemographic variables. Pearson’s Independent Chi-square test suggested that at 95% Confidence Interval (CI), gender, age, education, region, religion, ethnicity, profession, and income independently associated with rat meat consumption in Ghana (Table 3). However, no significant association exist between marital status, employment, and residential status with rat meat consumption (Table 3).

**Table 3.** Sociodemographic variables and rat meat consumption.

| Sociodemographic variables | Consume rat meat |  | Chi-square test (95%CI) |
| --- | --- | --- | --- |
|  | Yes (%) | No (%) |  |
| **Gender** |  |  |  |
| Male | 196 (23.8) | 298 (36.1) | $\chi^2=11.1$; $p=0.01$; $df=1$ |
| Female | 94 (11.4) | 237 (28.7) |  |
| **Marital Status** |  |  |  |
| Single | 163 (19.8) | 277 (33.6) | $\chi^2=4.7$; $p=0.10$; $df=2$ |
| Married | 103 (12.5) | 227 (27.5) |  |
| Married in the past | 28(3.4) | 31(3.7) |  |
| **Age** |  |  |  |
| <21 | 15 (1.8) | 48 (5.8) | $\chi^2=15.3$; $p=0.01$; $df=5$ |
| 21-26 | 70 (8.5) | 175 (21.2) |  |
| 27-34 | 95 (11.5) | 136 (16.5) |  |
| 35-44 | 62 (7.5) | 114 (13.8) |  |
| 45-54 | 34 (4.1) | 44 (5.3) |  |
| 55+ | 14 (1.7) | 18 (2.2) |  |
| **Education level** |  |  |  |
| No formal edu. /Primary | 68 (8.2) | 130 (15.8) | $\chi^2=20.9$; $p=0.000$; $df=3$ |
| Junior High School | 55 (6.7) | 46 (5.6) |  |
| Senior High School | 110 (13.3) | 215 (26.1) |  |
| Bachelor and above | 57 (6.9) | 144 (17.5) |  |
| **Regional division** |  |  |  |
| Northern Ghana | 57 (6.91) | 226 (27.4) | $\chi^2=42.6$; $p=0.000$; $df=1$ |
| Southern Ghana | 233 (28.2) | 309 (37.5) |  |
| **Religion** |  |  |  |
| Christianity | 183 (22.9) | 259 (32.4) | $\chi^2=34.6$; $p=0.000$; $df=2$ |
| Islamic | 80 (10.0) | 244 (30.5) |  |
| Traditionalist/Atheist/Others | 21 (1.5) | 12 (2.6) |  |
| **Ethnicity** |  |  |  |
| Mole-Dagbon | 51 (6.4) | 200 (25.0) | $\chi^2=61$; $p=0.000$;<br>df=9 |
| Akan | 102 (12.8) | 149 (18.6) |  |
| Ga | 39 (4.9) | 65 (8.1) |  |
| Ewes | 47 (5.9) | 34 (4.3) |  |
| Mamprusi | 15 (1.9) | 7 (0.9) |  |
| Dagati | 6 (0.8) | 15 (1.9) |  |
| Frafra | 3 (0.4) | 8 (1) |  |
| Guan | 4 (0.5) | 2 (0.25) |  |
| Others | 17 (2.1) | 36 (4.5) |  |
| **Profession** | | | $\chi^2=9.8$; $p=0.02$;<br>df=3 |
| Students | 53 (6.4) | 150 (18.2) |  |
| Health Professionals/Teachers | 64 (7.8) | 100 (12.1) |  |
| Traders | 83 (10.1) | 140 (17) |  |
| Others | 90 (10.9) | 145 (17.6) |  |
| **Employment status** | | | $\chi^2=1.3$; $p=0.26$;<br>df=1 |
| Employed | 161 (19.5) | 275 (33.3) |  |
| Not Employed/Retired | 129 (15.6) | 260 (31.5) |  |
| **Income (GHC.)** | | | $\chi^2=10.3$; $p=0.02$;<br>df=2 |
| <500/No income | 104 (12.6) | 150 (18.2) |  |
| 600-1100 | 64 (7.8) | 167 (20.2) |  |
| 1200+ | 56 (6.8) | 89 (10.8) |  |
| **Residential status** | | | $\chi^2=7.2$; $p=0.07$;<br>df=2 |
| Own the home | 57 (6.9) | 98 (11.9) |  |
| Family home | 138 (16.7) | 308 (36.6) |  |
| Renting | 90 (10.9) | 125 (15.2) |  |

### Logistic regression analysis of sociodemographic predictors and rat meat consumption

Bivariate and multivariable odd ratios from logistics regression models regressing sociodemographic variables (independent variables) with rat meat consumption variables unveiled significant associations. In the bivariate analysis, males exhibited 3.3 and 1.7 times higher odds of consuming rodent meat (OR=3.34; 95% CI: 2.47-4.50) and rat meat (OR=1.67; 95% CI: 1.23-2.23) respectively, compared to females. Participants aged 27-34years had 2.3 and 1.8 times higher odds of consuming rodent meat (OR=2.3; 95% CI: 1-62-3.32) and rat meat (OR=1.83; 95% CI: 1.28-2.63) respectively, compared to participants less than 26 years old. Participants with an age 35 years and above (35+) had 4.7 times and 1.6 times higher odds of consuming rodent meat (OR=4.676; 95%CI: 3.226-6.779) and rat meat (OR=1.64;95% CI:1.16-2.32) respectively, than those with age 26 years and below. Compared to Christians, Muslims had 1.4 times higher odds of consuming rodent meat (OR=1.45; 95% CI: 1.07-1.97). On the contrary, Muslims had 0.46 times lower odds of eating rat meat than Christians (OR=0.46; 95% CI: 0.34-0.64). Compared to the Akans ethnic group, Mole-Dagbon (OR=2.06; 95% CI: 1.42-3.1) and others (OR=1.75; 95% CI: 1.23-2.48), had high odds of eating rodent meat. On the contrary, Mole-Dagbon had 0.4 times lower odds of eating rat meat compared to the Akans (OR=0.37; 95% CI: 0.25-0.56). Regarding professions, health professionals/teachers (0.53; 95%CI: 0.34-0.82), and students (OR=0.28; 95%CI: 0.19-0.43), had lower odds of consuming rodent meat than traders. Students had 0.6 times lower odds of eating rat meat than traders (OR=O.6; 95% CI: 0.39-0.90). Compared with those receiving monthly income of 600 (GHS.) and above, those with 500(GH.) and below (including those with no monthly income) had 1.8 times higher odds of eating rat meat (OR=1.81; 95%CI: 1.24-2.65). With reference to region, Southerners had 0.6 times lower odds of consuming rodents than the Northerners (OR=0.63; 95%CI: 0.46-0.86). On the contrary, southerners had three times higher odds of consuming rat meat than Northerners (OR=2.99; 95%CI: 2.14-4.19). Compared to those below senior high school education (<SHS)/basic education/no formal education those to Bachelor degree, the latter ground had a 0.6 and 0.4 times lower odds of consuming rodent (OR=0.39; 95% CI: 0.27-0.57) and rat meat (OR=0.57; 95% CI: 0.39-0.83) respectively. Those with SHS education had 0.59 times less odds of eating rodent meat than those below SHS education (OR=0.59; 95% CI: 0.42-0.83). A strong association existed between those who consume both rodent and rat meat (χ=148; p=0.000; df =3). Multivariable binary regression model adjusting for other sociodemographic variables including those with p≤0.25 during the bivariate test suggested that being a male between 27-34yrs. of age, with no basic education/no formal education in the southern part of the country is most likely to consume rat meat (Table 4). The Table 4 contains only sociodemographic variables that showed significance in the adjusted model.

**Table 4.**
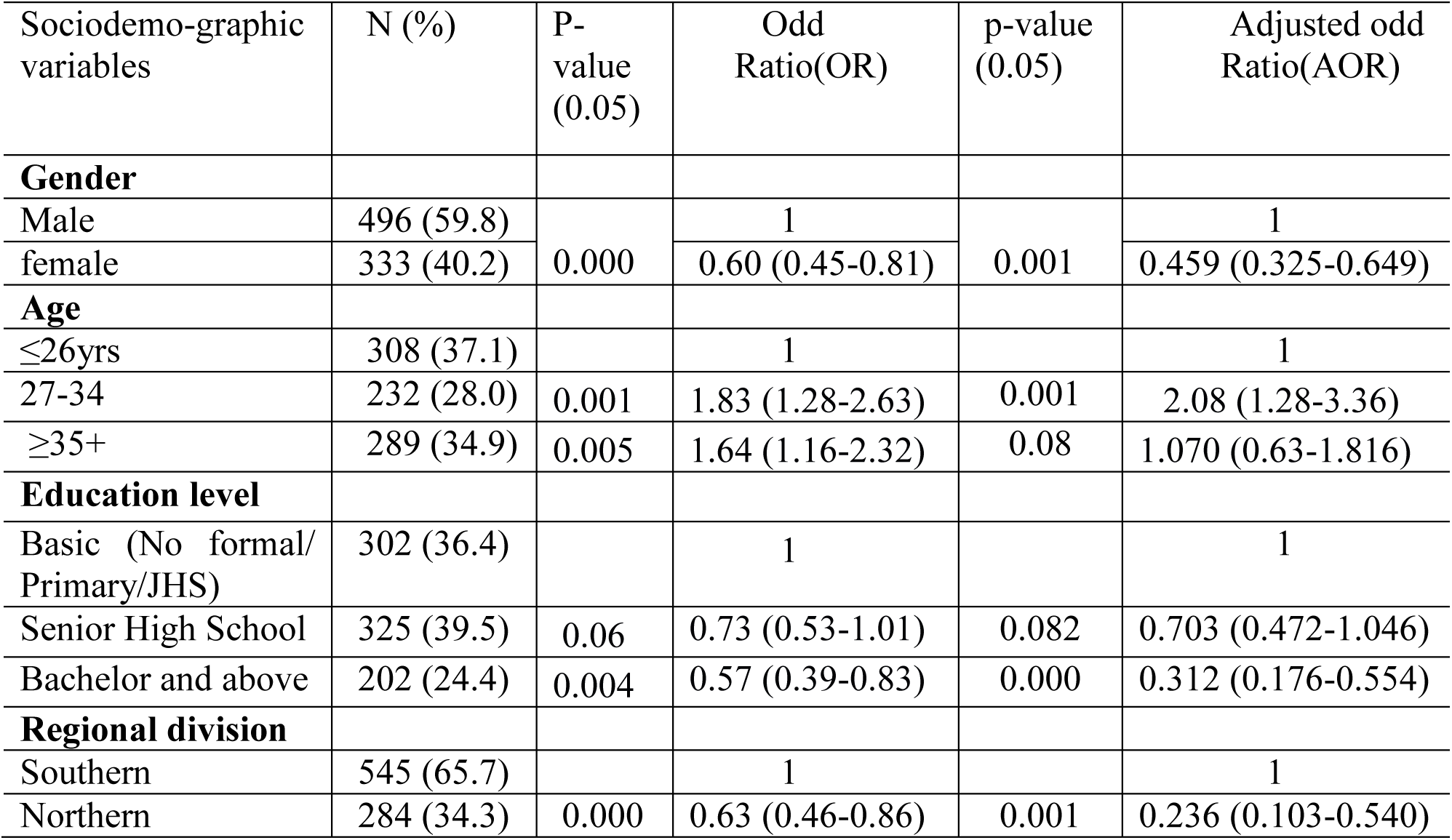
Multivariable logistics regression analysis of sociodemographic predictors and rat meat consumption in the participants.

### Perception of disease risks associated with rats

The results in Table 5 indicates the perception of disease risk associated with rat meat consumption. The findings generally indicate that, more than half of the study participants had very low to low perception scores on disease risks associated with rat meat consumption. Of the 829 participants, more than half (disagreed/strong disagreed=54.8%) indicated that rats found in the bush were different from rats found in towns whereas around 17% were indeterminate (neutral). A Significant number of participants (disagreed/strongly disagreed=61.4%) thought there were no disease risks associated with consuming rats from the bush and that rats from such locations did not carry disease agents, whereas, 13.5% had a neutral perception on the issue. About 38% of the participants disagreed that they could contract diseases by consuming rats in their neighborhood which was around half of the participants who indicated that rats from the bush cannot carry disease agents. Around 27% of the respondents had a neutral perception about contracting diseases from consuming town rats. This constituted twice the number of those who had a neutral perception of the disease agents’ status of bush rats (Table 5). Participants had low perception scores on rat-associated health risks in their neighbourhood (Table 5) primarily due to limited knowledge on rat-borne zoonoses. A significant majority (74.9%) perceived that rats in their neighbourhood posed a less severe (28%) to no health risks (46.9%). When asked if rats in their neighbourhood may transmit/carry any disease agents, 41% of the participants disagreed, whereas, 14.2% were neutral. On average 93% had no knowledge of anyone that had died or fallen sick due to rat meat consumption

**Table 5.** Participants’ perception of disease risks concerning rat meat consumption and rat infestation in neighbourhood.

| Statement/question | Response (%) |
| --- | --- |
| **I. I do not consider any difference between bush rat and town rat.** |  |
| Agree strongly | 74 (8.9) |
| Agree | 159 (19.2) |
| Neutral | 142 (17.1) |
| Disagree | 382 (46.1) |
| Disagree strongly | 72 (8.7) |
| **II. You can contract diseases by consuming rat killed from the bush** |  |
| Agree strongly | 60 (7.2) |
| Agree | 147 (17.7) |
| Neutral | 112 (13.5) |
| Disagree | 429 (51.7) |
| Strongly disagree | 80 (9.7) |
| **III. You can contract diseases by consuming rat killed from the neighbourhood** |  |
| Agree strongly | 64 (7.7) |
| Agree | 224 (27.0) |
| Neutral | 224 (27.0) |
| Disagree | 246 (29.7) |
| Strongly disagree | 70 (8.4) |
| **IV. know someone that has fallen sick from eating rat meat** | No=779 (94.0) |
| **V. Know someone that has died from eating rat meat.** | No=762 (92.0), |
| **I. Rats in my neighbourhood have:** |  |
| highly severe health risk | 208 (25.1) |
| Less severe health risk | 232 (28) |
| No health risk | 389 (46.9) |
| **II. Rats in your neighbourhood may carry disease agents/transmit disease** |  |
| Agree strongly | 126 (15.2) |
| Agree | 241 (29.1) |
| Neutral | 118 (14.2) |
| Disagree | 311 (37.5) |
| Strongly disagree | 33 (4.0) |

### Sociodemographic predictors of perception scores on disease risk associated with rats

Mann Whitney U test results detailed in Table 6 suggested a significant proportional difference in mean perception scores on health risks associated with rat meat consumption between the genders. No significant difference exist between southern and northern Ghana in mean perception scores on disease risks associated with rat meat consumption. More males than females considered there was a difference between town and bush rats. Both males and females equally indicated that diseases can be contracted by consuming rats from their neighbourhood, however, more males than females perceive a disease risk with consuming bush rats. There was no significant difference between northerners and southerners perceptions of the disease risks pertaining to consuming rat killed from the bush or town. However, a high proportion of northerners than southerners perceive a difference between town and bush rat (Mann Whitney U= 99367; p=0.000). A significant proportion of participants that consumed rat meat rejected the assumption that they can contract diseases by consuming town rat meat (Whitney U=67956.0; p=0.003). They significantly disagreed that rats in their neighbourhood may carry disease agents (Mann Whitney U= 60101; p=000). However no significant difference exist in their perception of disease risk on consuming bush rat (Whitney U=78542.0; p= 0.675). Kruskal-Wallis test of independence and pairwise post hoc tests adjusted by Bonferroni correction for multiple tests detailed in Fig 2 suggested a significant proportional differences in mean perception scores on disease risks associated with consuming neighbourhood/town rats between the education levels, ethnic and professional groupings. Akan ethnicity, those with bachelors’ education and above, and health professionals/teachers, had significantly high mean risk perception scores compared with their counterparts. However, no significant difference existed in the mean risk perception scores among the religious groups. A one way ANOVA showed no significant difference among the age groups regarding their perceptions that diseases can be contracted through the consumption of bush rat meat (F=0.69;p=0.51; df=2) or town rat meat (F=0.21;p=0.81;df=2)

**Fig 2:**
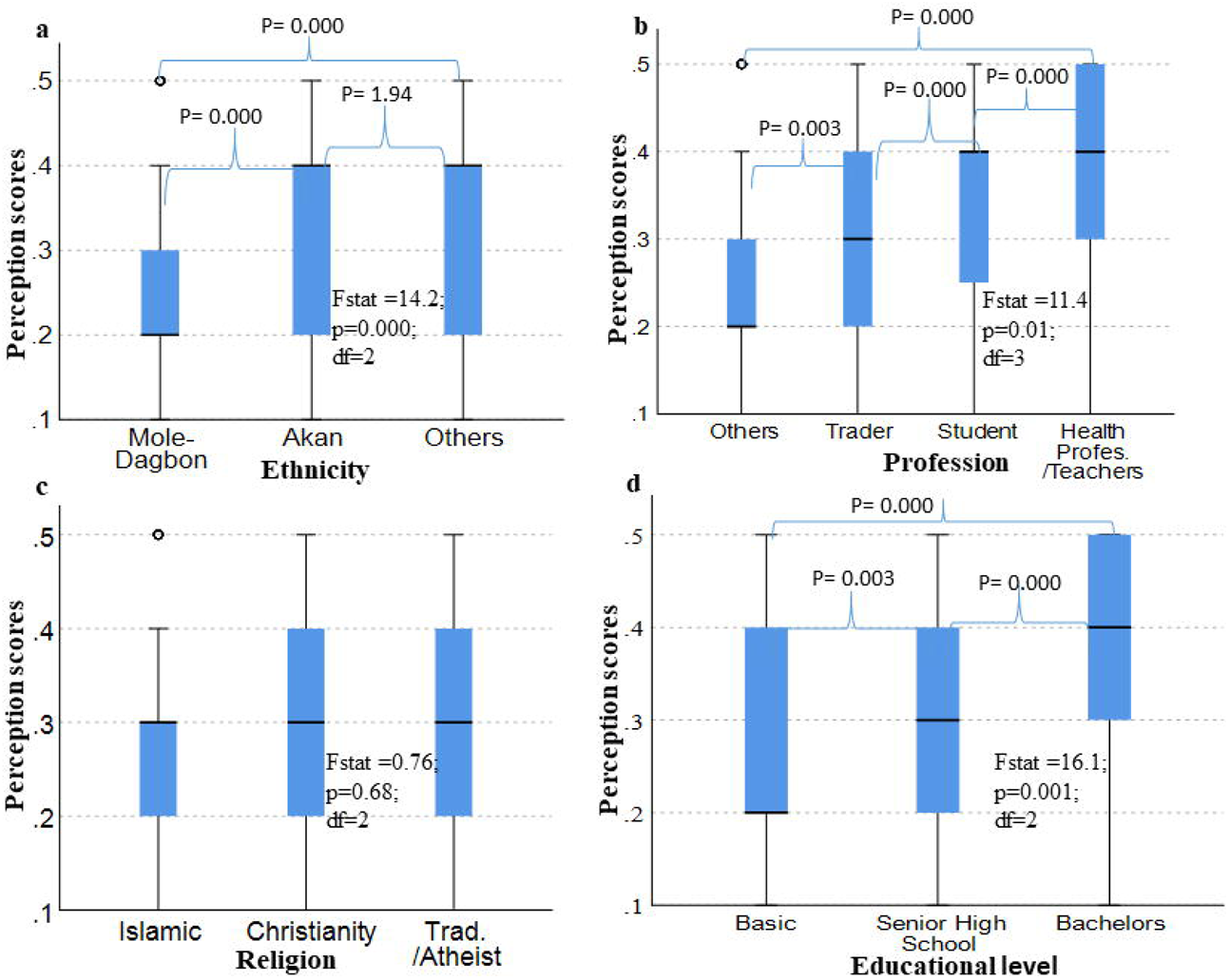
Correlation between respondents’ sociodemographic profile and perception scores on disease risks associated with consuming neighbourhood rats. Ethnicity (a), Profession (b), Religion (c), Education (d)

**Table 6.** Mann Whitney U test analysis of correlation between sociodemographic variables and perception scores on disease risks associated with rat meat consumption [95%CI].

| Socio.<br>variables | Statement/question: Perceptions about disease associated with rats |  |  |
| --- | --- | --- | --- |
|  | I. Considers difference between bush and town rat. | II. Can contract diseases by eating rat killed from the bush. | III. Can Contract diseases by eating rat killed from neighbourhood/town |
| **Gender** | U = 69009; z-score= 4.3<br>p=0.000 | U 76567.5;z-score = -<br>2.1; p=0.033 | U=84266;z-score=0.57;<br>p=0.57 |
| Male: mean rank | 442.1 | 426.3 | 410.8 |
| Female: mean rank | 374.2 | 396.9 | 420.5 |
| **Region** | U=99367; p=0.000; z-score =7.13 | U=73208;p=0.13; z-score= -1.5 | U=71099.0; p=0.051; z-score= -1.9 |
| Southern: mean rank | 374.7 | 421.9 | 425.8 |
| Northern: mean rank | 492.4 | 400.3 | 392.9 |

Males, basic education leavers, those from Mole-Dagbon ethnicity, traders, those who are Islamic/Muslim, and reside in northern Ghana proportionally showed low perception scores about disease risk associated with rats in their neighborhood (Table 6 and Fig 3). Between the genders, a significant proportion of females than males perceived a disease risks associated with rats in their neighbourhood (U = 95867; P=0.000; z-score=4.1). More southerners than northerners perceive disease risks associated with rats in their neighbourhood (U=47887.0; p=0.00; z-score = -9.4). Kruskal-Wallis test of independence and pairwise post hoc tests adjusted by Bonferroni correction for multiple tests suggested a significant proportional differences in mean perception scores on disease risks associated with rats in neighbourhood among the religious groups, educational levels, ethnic and profession groupings (Fig 3). Christians compared with Islamic/Muslims have a higher perception of the disease risk associated with rats in their neighbourhood (Fig 3). Mean perception scores increased significantly from a low perception score in basic education leavers to intermediate/neutral perception in senior high school to a higher perception score in bachelor education and above (Fig 3). Among the professions, health professionals/teachers and students obtained higher mean perception scores than traders. Among the ethnic groups, Akans had a higher perception score than the Mole-Dagbon.

**Fig 3.**
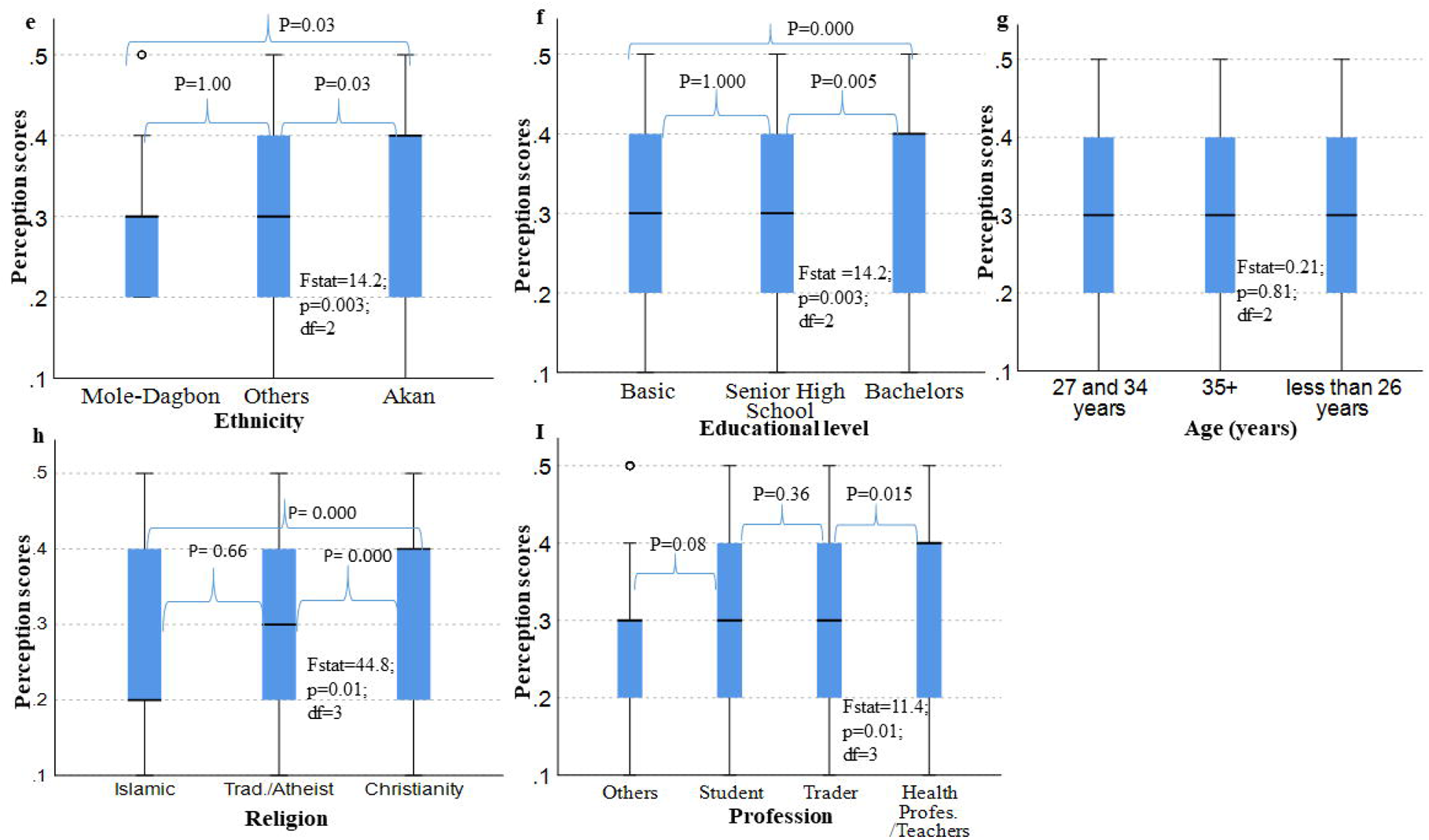
Correlation between respondents’ sociodemographic profile and perception scores on neighborhood rats as carrier of disease agents. Ethnicity (e), Educational level (f), Age (g), Religion (h), Profession (I)

### Survey respondents’ awareness about rat infestation

This section presents a summary on respondents’ awareness about rat population increases in their neighbourhood (Table 7). In general, participants had high awareness level of rat infestations in their neighborhood. Of the 829 study participants, a significantly high number (35.7%) of them confirmed seeing rats daily. About 11.9% of the participants sighted rats weekly and 13.6% and 31.1% sighted rats in their neighborhood on monthly and occasional basis respectively (Table 7). About 17.9% of the survey respondents believe that rats’ population has increased in their neighborhood whereas 24.5% observed that the rats are already common within their neighborhood (Table 7). Regarding factors that accounted for rats’ infestations, half of the participants have no knowledge of the factors accounting for the presence of rats in their neighborhood. Less than 20% of the participants attributed the rodents to poor drainage systems, change in weather conditions, poor sanitation and poor housing (Table 7). A significantly high number of the participants (60.3%) indicated rats are mostly found in houses and community drains and more than half of the participants know a place around and in their residence where rats are presently found. Some 19.9% indicated that rats often visit their homes (Table 7).

**Table 7.** Respondents’ awareness about rat infestations in their neighbourhood.

| Statement/Question | Response (%) |
| --- | --- |
| **I. How often do you see rats in your neighbourhood?/rat sighting frequency** |  |
| Daily | 295 (35.7%) |
| weekly | 99 (11.9) |
| monthly | 113 (13.6) |
| Occasionally | 258 (31.1) |
| Can't recall | 34 (3.8) |
| **II. Do you observe that rat population in your neighbourhood has increased?** |  |
| I haven't noticed | 473 (57.1) |
| I have observed that | 148 (17.9) |
| Already common and haven't noted increase | 203 (24.5) |
| **III. Do you have any idea on what can account for rats' prevalence in your neighbourhood?** |  |
| No idea | 416 (50.2) |
| Poor drainage system | 14 (1.7) |
| Change in weather condition | 44 (5.3) |
| Poor sanitation/filthy Environment | 64 (7.7) |
| Poor housing structure | 37 (4.5) |
| **IV. Know a place in the home presently where rats are.** | Yes = 465(56.1) |
| **V. If no, do they visit the home occasionally** | Yes 165(19.9) |
| **VI. In which season do you commonly find rats in your neighbourhood?** |  |
| Both | 444 (53.6) |
| Dry | 283 (34.1) |
| Rainy | 72 (8.7) |
| **VII. Where are rats mostly found in your neighbourhood?** |  |
| Houses | 286 (34.5) |
| Community drains | 214 (25.8) |
| Public places | 65 (7.8) |
| Others | 110 (13.3) |
| Can't tell | 154 (18.6) |

### Survey participants’ concerns about rats in their neighbourhood

Table 8 indicates the results of the 829 survey participants concerns about rats in their neighbourhood. Although a significant majority of the respondents (70%) were concerned about rats in their neighborhood/home, the majority made no attempt (56%) to control the rats. Of those concerned about rats, the highest number of them were concerned about the economic damage rats’ cause to properties and food products (41%). Others were concerned about the disease/health risk (18.0%) and public nuisance (10.4%). Around 28.2% were not concerned about rats at all.. When finally asked what participants will do when rats enter their homes, half the survey participants will attempt to kill the rat and dispose of it, 24% will kill the rat for food, 4.1% will contract someone to kill the rat, and a high number (16.9%) will do nothing.

**Table 8.**
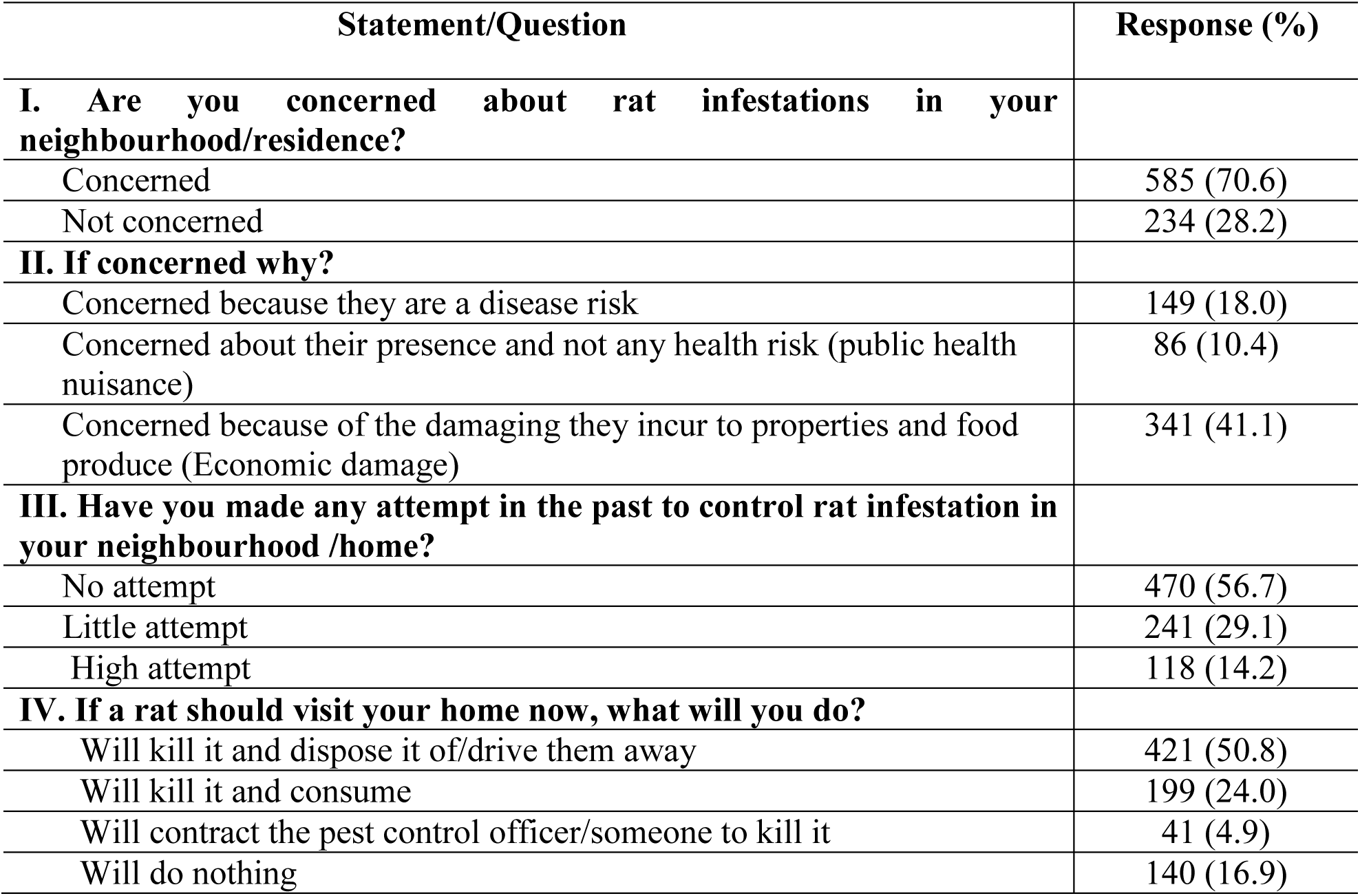
Participants’ concerns about rat infestation in their neighbourhood.

## Discussion

This survey presents the first baseline study on urban residents’ interactions and attitudes towards rodents in Ghana, which is particularly pertinent given the recent emergence of Lassa fever in the country, and the widespread occurrence of drug resistant pathogens. Our research has identified weaknesses in community knowledge of rodents and the diseases they may spread. Hopefully our data contribute to the on-going efforts by the Ghana Health Service to minimize these health problems through targeted educational campaigns aimed at those groups that where knowledge gaps were identified. Proportionally, our study population was predominantly represented by the two ethnic majorities in Ghana, the Akans and the Mole-Dagbon that collectively constitute 63% of Ghana’s population, which is why approximately 61% of our study subjects were from these ethnic categories. The Akans are regionally prevalent in southern Ghana whereas, the Mole-Dagbons are widespread in the North, as well as Muslim religious groups. We received a higher number of people originating from southern Ghana (65.7%) compared to north, which was attributed to the greater urbanization and population density in the south.

The present study delved into rat meat consumption patterns and perception of disease risks pertaining to peridomestic rats. Sociodemographic determinants associated with the risk of rodent-borne diseases were also assessed. In summary, our findings revealed that rats including commensal species are consumed by a considerable number of Ghanaians in urban settings. Both rat meat consumption and perception of disease risks were driven by sociodemographic variables. Overall statistical analyses showed that, except for education and profession that showed some positive influence on the practice of rat meat consumption and perception of risks, rat meat consumption did not correlate with perceive risk in most sociodemographic categories. The perception of risks about rat meat consumption and rat infestation in urban environments were generally low among the participants allowing us to reject our hypothesis that urban dwellers in Ghana have high risks perceptions pertaining to peridomestic rats. We surmised that this low perception may have contributed to the limited efforts made by the public to control rats.

Our literature search revealed a publication scarcity on the consumption rate of rats in various country settings and regions. Previous research primarily focused on either rodent or bush meat consumption in general, therefore our findings were discussed in that regard. In our study, approximately 65% consumed rodents. This finding aligns with recent studies in Ghana indicating that 67% of Ghanaians [28] prefer bush meat, including rodents, due to the flavor and taste. However, the rate of rodent consumption was higher in Guinea, West Africa, where a study showed that 91.5% of the population considers rodents as an important protein source [24] and also that they are common and easy to obtain at mere/no cost implication. In the present research, we found that approximately 35% of rodent meat consumers also consumed rat meat, and 26% will consume commensal rat in their neighbourhood when available. These findings closely coincide with a related study in Nigeria, which found 33.5% and 31.1% of the study respondents interviewed from urban and peri-urban neighborhood respectively consume urban rodents [23].

Preferences for consuming wild meats in Ghana and across Africa are influenced by various sociodemographic factors [57,62,63]. In this study, we identified gender, education, sex, age, religion, ethnicity, region, income, and profession as independently associated with rat meat consumption. Rat meat consumption correlated with religious beliefs with higher consumption rates in Christians than Muslims aligning with the report by [28] which could be attributed to their religious beliefs. Additionally, cultural beliefs among different ethnic communities contribute to varying levels of bush rat meat consumption. Particularly, high rat meat consumption was observed among the Akan ethnicity as observed by [59]. Although limited data were available for the Ewes and the Mamprusi ethnicities, our data indicates that there is a higher proportion of rat meat consumers within this group. The influence of ethnicity on bush meat consumption has been well-documented in the literature [57,62,63,64]. Regarding regional disparities, our study revealed, a lower consumption rate among the northerners compared to the southerners which was attributed to a higher number of Muslims in the region particularly among Mole-Dagbon ethnic group who probably appear to exhibit less preference for rat meat, presumably due to their beliefs. Gender imbalances in wild meat consumption, with females exhibiting reduced preferences were previously evidenced in Ghana [57]. Regarding age and educational related disparities, our findings are consistent with related research indicating that individuals with at least senior high school education demonstrated reduced wild meat consumption [57]. Higher educational levels have been associated with decreased hunting and wild meat eating behaviours [57]. Regarding income, there are prevailing arguments on if wild meats are a luxury or necessary survival food for the poor [65]. In our research, participants with low or no income associated strongly with rat meat consumption, especially males aged 27 to 34years with no formal/basic education in southern Ghana. The monthly minimum wage in Ghana is only around 500 cedis (USD$37.0 equivalence) suggesting that there is a correlation between rat meat consumption and poverty or low living standards. This finding aligns with the research by [66]. While previous study found no relationship between bush meat consumption and profession [66] the present study did find such an association. Health workers/teachers and students consumed less rat meat than traders, attributable to the high perception scores of disease risks in those with a higher education. We also found that education, profession, religion, and ethnic disparities produced significance differences between the respondents regarding the disease risks associated with peridomestic rats. Male genders, Traders, those with a basic education, Muslims, the Mole-Dagbon ethnic group, and northerners obtained lower perception scores on the disease risks associated with rats in their environment. Regarding the regional distribution of perception scores regarding commensal rats, proportionally, higher perception scores were recorded in southern Ghana than the north. As highlighted earlier, there are more Muslims widespread in the north of Ghana and among the Mole-Dagbon ethnic group who have less preference for rat meat, yet still had low perception scores on disease risk of rodent-borne diseases. Southern Ghana encompasses the major cities of the country and is highly urbanized, with higher levels of education. As noted, other studies have found that education is a strong determinant for knowledge on animal related disease risks [57,67]. In a related study, perception risk scores varied regionally, and by education; rural based cities obtained lower perception scores than urban areas and those with higher education had high risk perception scores than those with a basic education [57]. This again demonstrates that education is a strong determinant for risk perception hence, education and awareness programs are an important intervention strategy to enhance knowledge and improve perceptional awareness on rat-borne diseases. In our study, there were some notable discrepancies in the perception scores between the genders, with males having a lower perception score on the health risks and disease transmission risks associated with commensal rats. This is probably why men tend to consume rat meat more often than the females. In the study herein participants showed significantly high concerns about rat infestation in their neighborhood mainly due to the economic hazard caused by rats such as damage to food and property. Despite their concerns, limited attempts were made to manage rats in their residence/neighbourhood. The limited control attempts can be attributed to the low prevalence of the disease risks associated with rats.

In our current investigation, 35% of the participant reported a daily encounter with commensal rats. Considering the reported weekly, monthly and occasional sightings. A rise in rodent numbers can be attributed to shifts in human environment, ecology and climate [2–20] with research indicating a heightened risk of rat infestation in neighborhoods characterized by high poverty levels, aged buildings, dense housing systems and inadequate sanitation [11,20,21]. Similar to most other African nations, Ghana has a persisting sanitation challenges and poor drainage management contributing to the proliferation and establishment of pest populations.

Despite a significant proportion of participant Ghanaian’s across the various sociodemographic categories interact seeing rats in their neighborhood and that a considerable number consume them, the perceptional awareness on the disease risks regarding rats is low. It is probable that the lack of education and awareness programs pertaining to rat-borne zoonoses in Ghana and the rare occurrences of rat-related zoonoses in the past is the cause for the lack of awareness of the current disease risks. Over 93% of the study participants have no knowledge of anyone suffering from any rat borne zoonoses or have died from rat meat consumption, neither had they seen any information on this topic from the media. Poor perceptional awareness on disease risks associated with human interaction with rats has been previously noted across Africa and Asia [36,55,56,57,58]. A study conducted in rural Ghana documented low awareness level among rural dwellers regarding the potential risks of consuming wild meats [56]. Similarly, in a related study on the consumption of fruit bats in Ghana, although consumption of these species was high among the locals, participants held little beliefs on disease risks [57]. In Malaysia, despite a high proportion of respondents consuming bush meat, the majority had minimal knowledge of the disease risks [58]. Likewise, in Tanzania, more than half the proportion of local population were unaware of the disease risks relating to handling bush meats [55]. The significant majority of the study participants in our investigations found no disease risks associated with consuming rats from the bush compared to rats from town. This aligns with a related study in Uganda, where cooks of bushmeat considered common edible bush rat meat consumption as the least likely to result in any sickness [34]. Among pastoralists in northern and eastern Tanzania, there was a widespread of skepticism that zoonotic illnesses could be spread through the consumption of animal products [36]. We found that the proportion of participants willing to buy bush rat or town rat meat sold in a restaurant were similar suggesting that rat meat consumers consume both commensal and bush rat. People interacting with rats’ may inhale aerosolized pathogens from the fur of rats, can contract blood-borne infections during the handling and butchering process of the carcasses, and can be exposed to pathogens through the consumption of undercooked rat meats [24,34,36]. Mustomys rats that transmit Lassa Fever are highly prolific and commensal in West, Central and East Africa inhabiting areas that are overcrowded and unsanitary [43,44,68] enhancing tendency of being consumed. In Nigeria, the over 85% of the study respondents who had no knowledge of LF and disputed the existence of Lassa Fever virus in rats consume the *Mastomys* rat vectors [23] exposing them to disease risks. In our investigation, the majority of our respondents indicated rats are most present in their homes and community drains. In the previous study, 28% of households who had rodents in their residence come into contact with their urine and droppings and 24% eat rodent contaminated foods [23]. Human interactions with rats in the home provides a pathway of transferring zoonotic illnesses into the human population via direct and indirect route [32]. Contact with rats through scratches or bites, and exposure to their secretions constitute direct transmission route [3,69]. Hantavirus Pulmonary Syndrome is contracted via direct transmission through breathing in aerosolized secretions from rats [70]. Indirectly, rats could transmit diseases to human through stepping on household water sources, food, clothing and floors [71] and exposure to air-bone diseases from rats through inhaling sweeping dust can also occur [72]. Peridomestic rats are sentinels, reservoir and vectors in the transmission of AMR pathogens to humans and other animals since they share similar environment with people, domestic pets and insect vectors [47]. The inhabiting of diverse habitats including municipal waste water systems, livestock, urban agriculture sites, cemeteries, mortuaries and biomedical/healthcare/hospital wastes disposal sites, makes urban rats’ good candidates in the transmission of a wide range of AMR [47]. These pathogens may be spread to homes, and domestic animal populations [48] with widespread reports of multiple drug resistant pathogens being spread by commensal rats [31,32,49,50,51]. Rats are also responsible for nosocomial infection transfer [73], and homes close to healthcare and cemeteries in urban areas may be at a higher risk of infections. Rats cause cross food contamination in homes and restaurants by transferring pathogens and chemicals from the various sites to cooked food and stored products [74]. The detrimental implications associated with rats make their presence in urban settlements a critical health risk.

## Conclusion

In summary, this study investigated rat meat consumers among urban Ghanaians and examined the perception of risk in the consumption of rat meat and the presence of rat infestations. The study characterized the sociodemographic determinants of rat meat intake and the perceived risk associated with rats. The findings revealed that a considerable proportion of the Ghanaian population in urban areas consume rats, including commensal species. Risk perceptions relating to disease risk associated with rat meat consumption and peridomestic rats were generally low hence, limited attempts were made to control rodents in their neighborhood. The lack of public education and awareness programs on rat-borne zoonoses may be a contributing factor for the low risk perception. Sociodemographic determinants of rat meat consumption and risk perception were identified as targets for future interventions. Our findings reveal that Ghanaians in urban areas are at a threat of zoonotic spillover, which highlights the critical need for public education on rat-borne zoonoses. In addition, rodent management in urban Ghana requires an immediate prioritization, as rodents are a source of drug resistant pathogens that can be spread through communities. Our study will be useful to the Ghana Health Service in the on-going measures to prevent Lassa fever re-occurrence and the spread of drug resistant pathogens in urban areas.

## Data Availability

All data produced in the present work are contained in the manuscript

## Data Availability

All data produced in the present work are contained in the manuscript

## Acknowledgement

We acknowledge the support of executives of the Ghana Environmental Health Officers Association, health officers and trainees from the Accra School of Hygiene from the various administrative regions in Ghana. We especially wish to thank these trainees, Miss Matilda Owusuaa from the Bono region of Ghana, Mr. Alhassan Baako Abdullai from Yendi, Northern Ghana and Mr. Bentum Gabriel for their efforts.

